# A Survey of Individuals’ Willingness to Share Real-World Data Post-Mortem with Researchers

**DOI:** 10.1101/2024.05.13.24307263

**Authors:** Rachele M. Hendricks-Sturrup, Christine Y. Lu

**Affiliations:** Department of Population Medicine, Harvard Pilgrim Health Care Institute and Harvard Medical School, Boston, MA, USA; Duke-Robert J. Margolis, MD, Institute for Health Policy, Washington, DC, USA; Department of Interdisciplinary Health Studies, Ohio University, Athens, OH, USA; Kolling Institute, Faculty of Medicine and Health, The University of Sydney and the Northern Sydney Local Health District, Sydney, NSW, Australia; School of Pharmacy, Faculty of Medicine and Health, The University of Sydney, Sydney, New South Wales, Australia

**Keywords:** data use, data privacy, data sharing, post-mortem, posthumous, medical websites, health data, real-world data

## Abstract

**Background:** Posthumous data use policy within the broader scope of navigating post-mortem data privacy is a procedurally complex landscape for those engaged in data collection and sharing, and data-driven health research.

**Objective:** To help address some of this complexity by exploring patterns in individuals’ willingness to donate data with health researchers after death and developing practical recommendations.

**Methods:** An electronic survey was conducted in April 2021 among adults (≥18 years of age) registered in ResearchMatch (www.researchmatch.org), a national health research registry. Descriptive and multinomial logistic regression analyses were conducted at a 95% confidence level to determine the association between willingness to donate data after death overall and across each demographic category (education level, age range, duration of using online medical websites, annual frequency of getting ill) and based on the overall quantity at which individuals expressed willingness to donate data (some, all, none) with researchers.

**Results:** Of 399 responses, most participants were willing to donate health data (electronic medical record data [67%], prescription history data [63%], genetic data [54%], and fitness tracker data [53%]) after death. We identified that individuals were more likely to donate some data after death (versus no data) if they had longer duration of using online medical websites (relative risk ratio = 1.22, p<0.05, 95% CI: 1.01 to 1.48). No additional significant findings were observed between willingness to donate all, some, or none of their data after death and other demographic factors.

**Conclusions:** There are opportunities to enhance practices to engage patients in health systems’ online medical websites as one potential mechanism to encourage or inspire individuals in posthumous data donation for health research purposes.

## Introduction

Today, in the 21^st^ century, participation in the digital economy is not only commonplace, but also necessary for the majority of the population to engage in their day-to-day livelihoods. From searching the internet, using self-monitoring devices (i.e., wearables and fitness trackers), engaging on online banking, obtaining genetic ancestry test results, seeking medical care online, connecting on social media, online purchasing, ride-sharing, sending email and text messages, and more, the likelihood of data being stored in online accounts and individuals participating in the digital economy or development of digital assets at any age today is high. Therein is both a concept and practice of data ownership and individual autonomy to donate data for various causes or initiatives, such as health research, during life and/or death. Those who make decisions regarding the collection and subsequent uses of individuals’ data upon their death, or posthumous data use within the broader scope of post-mortem data privacy and discretion, must navigate a procedurally complex landscape.^1–5^

For instance, decision-makers must consider existing and applicable data privacy law, review available decisions made in related cases that have been presented in arbitration or the court of law, and determine what is in the best interest and well-being of decedents’ surviving family members or life/care partners.^6^ Within the United States (US) the Health Insurance Portability and Accountability Act (HIPAA) protects individuals’ personally identifiable health information or data derived from health care settings for 50 years after death.^7,8^ Likewise, the European Convention of Human Rights also protects the confidentiality of persons’ medical information upon death.^9^ Yet, lessons learned from the legally contested commercialization of Henrietta Lacks’ immortal cancer cell line, an event that predates HIPAA, and use cases surrounding HIPAA limitations and other legal paradigms, or lack thereof, concerning data send an important message.^7,10,11^ That is, today’s general lack of comprehensive, or the current presence of outdated, posthumous biobank and data use consent practices for health research are a potential blind spot in data use policy today.

The amount of data related both directly and indirectly to health, and generated and/or stored within or outside of traditional health care settings, is unprecedented. Resulting are concerns about the sufficiency of data privacy and human subjects research protection laws, if any, to reach beyond enterprise terms of service agreements and protect against potential misuses of individual-level data of the deceased. Comprehensive laws such as the 2014 Uniform Fiduciary Access to Digital Assets Act (UFADAA; including its revision called RUFADAA) legally provides data subjects and their fiduciaries ownership and dictation rights over digital assets in the event a data subject or owner dies or becomes incapacitated.^12^ Yet, privacy law protections are limited or uneven at best. For instance, the European General Data Protection Regulation does not apply to deceased individuals and the new Washington My Health My Data Act in cases of deceased individuals.^13,14^ One legal scholar noted that the California Consumer Privacy Act “CCPA stops short of empowering the personal representatives of decedents to exercise control over personal data according to the decedent’s wishes.”^1^ Lastly, the US Common Rule does not consider research involving deceased individuals as human subjects research (must be a ‘living individual; 45 CFR 46.102[e1]) and thus does not require institutional review board oversight of research involving exclusively deceased individuals.^15^

A systematic review shows that individuals and their relatives, who might serve as personal representatives, are willing to share data with health researchers when given sufficient opportunities, within or outside of legal processes, to plan for and dictate uses of their data assets before becoming deceased.^16^ Further insights to help navigate this legally and procedurally complex frontier are necessary to drive innovative and trustworthy data-driven health research and reduce risk of data misuse for deceased individuals and their families, former caregivers, etc. In this paper, we report findings from a national survey eliciting individuals’ willingness to donate both the volume and type of data after death with health researchers and our analysis of how demographic factors might influence individuals’ willingness to donate data after death.

When considered alongside current and emerging evidence, policy, and commentary, our present findings will be useful to researchers, practitioners, and data subjects seeking to make immediate decisions about and/or collaborate on the development of policies, consent tools, and/or practices focused on posthumous data use within the broader scope of post-mortem data privacy.

## Methods

### Survey Development, Validation, and Distribution

The survey was developed, validated, and distributed in April 2021 and as described in detail in our prior work.^17,18^ The survey was made available to participants, who agreed to be contacted, using online Qualtrics software. Survey participants aged 18 years and older were identified and contacted online using the ResearchMatch online platform, a “disease-neutral, Web-based recruitment registry to help match individuals who wish to participate in clinical research studies with researchers actively searching for volunteers throughout the US.”

### Response Data Collection & Analysis

This analysis centers on participant responses based on four demographical items (independent variables) and two items focused on willingness to share sources of data with researchers (dependent variables). The present analysis centers on ResearchMatch participants with a 100% response or completion rate (inclusive of completed surveys with items containing no responses). A Qualtrics software tool was used to calculate an ideal survey sample size (n= 384) based on the total ResearchMatch population (95% CI; 5% margin of error). To observe the association between willingness to donate data after death and demographic variables, we fitted multinomial logistic regression models using each willingness to donate real-world data after death outcome as a dependent variable (no data, some data [willing to donate data from at least one source], or all data [willing to donate data from every source]). Independent variables were education level, age range, duration of using online medical websites, and annual frequency of getting ill (see Table 1). Results were reported as relative risk ratios (RRRs) with 95% confidence intervals. Descriptive analyses were conducted using Microsoft Excel and all logistic regression analyses were conducted using Stata version 17.0 Standard Edition.

**Table 1.** Demographic variables (age range, education level, duration of using online medical websites, annual frequency of getting ill) and willingness to donate specific real-world data sources after death.

| **Demographic Variables** |  |
| --- | --- |
| *Education Levels* | *Age Ranges (years)* |
| • High school<br>• Some College/Associates/Trade School<br>• Bachelors<br>• Masters<br>• Doctorate or other terminal degree | • 18 to 30 (Under 30)<br>• 31 to 40<br>• 41 to 50<br>• 51 to 60<br>• Over 60 |
| *Duration of Using Online Medical Websites (years)* | *Annual Frequency of Getting Ill* |
| • Never<br>• Less than 1<br>• 2 to 3<br>• 4 to 5<br>• 6 to 7<br>• More than 7 | • Less than 1<br>• 2 to 3<br>• 4 to 6<br>• 7 to 10<br>• More than 10 |
| **Real-World Data Sources (Willingness to Donate None, Some, or All)** |  |
| *Social media data*<br>• Facebook data<br>• Twitter data<br>• Instagram data<br>• Snapchat data<br>• Yelp reviews and ratings<br>*Health data*<br>• Fitness tracker data (FitBit, Apple Watch, etc.)<br>• Prescription history (CVS, Walgreen's, etc.)<br>• Electronic medical record data<br>• Genetic data (23andMe, etc.)<br>*Direct communication data*<br>• Text Message and Phone Call Data<br>• Email History (Gmail, Yahoo, Comcast, Verizon, etc.) | *Online browsing or streaming data*<br>• Music streaming data (Spotify, Pandora, etc.)<br>• Google search history<br>*Financial data*<br>• Online purchase history (Amazon, Target, Ebay, etc.)<br>• Tax records and income history<br>• Credit card statement data<br>*Location data*<br>• Ride-sharing history (Uber, Lyft, etc.)<br>• Geolocation (GPS from your phone or computer) data<br>*Voting history data* |

### Ethics Review, Oversight, and Approval

ResearchMatch is a registry and collaborative project that is maintained at Vanderbilt University and overseen by the Vanderbilt University Institutional Review Board. The present study was reviewed and approved by the Ohio University Institutional Review Board under protocol #20-E-457. ResearchMatch participants’ completion of the survey implied their consent to engage in the survey.

## Results

### Overall Participant Characteristics

Overall participant characteristics were as described in our prior work.^17,18^ Among 470 participants who initiated the survey, 402 completed and submitted the survey (86% completion rate). An overall majority of the of the survey participants were over the age of 21 (99%); held either some college/associates/trade school, a bachelors’ degree, or masters’ degree (87%); used online medical websites for 7 years or more (56%); and reported an annual frequency of getting ill of six occurrences or less (94%).

### Willingness to Donate Data After Death

Of the 399 participants who completed the survey, regardless of demographic details provided, 6.8% (n= 27) were willing to donate all data, 86% (n= 343) were willing to donate some data, and 7.3% (n= 29) were unwilling to donate no data after death. Most participants were willing to donate health data (electronic medical record data [67%], prescription history data [63%], genetic data [54%], and fitness tracker data [53%]) after death (see Figure 1; n= 399). Few participants were willing to donate Snapchat data (18%), credit card statement data (19%), tax records and income history data (21%), ride-sharing history (22%), and Twitter data (23%) after death.

**Figure 1.**
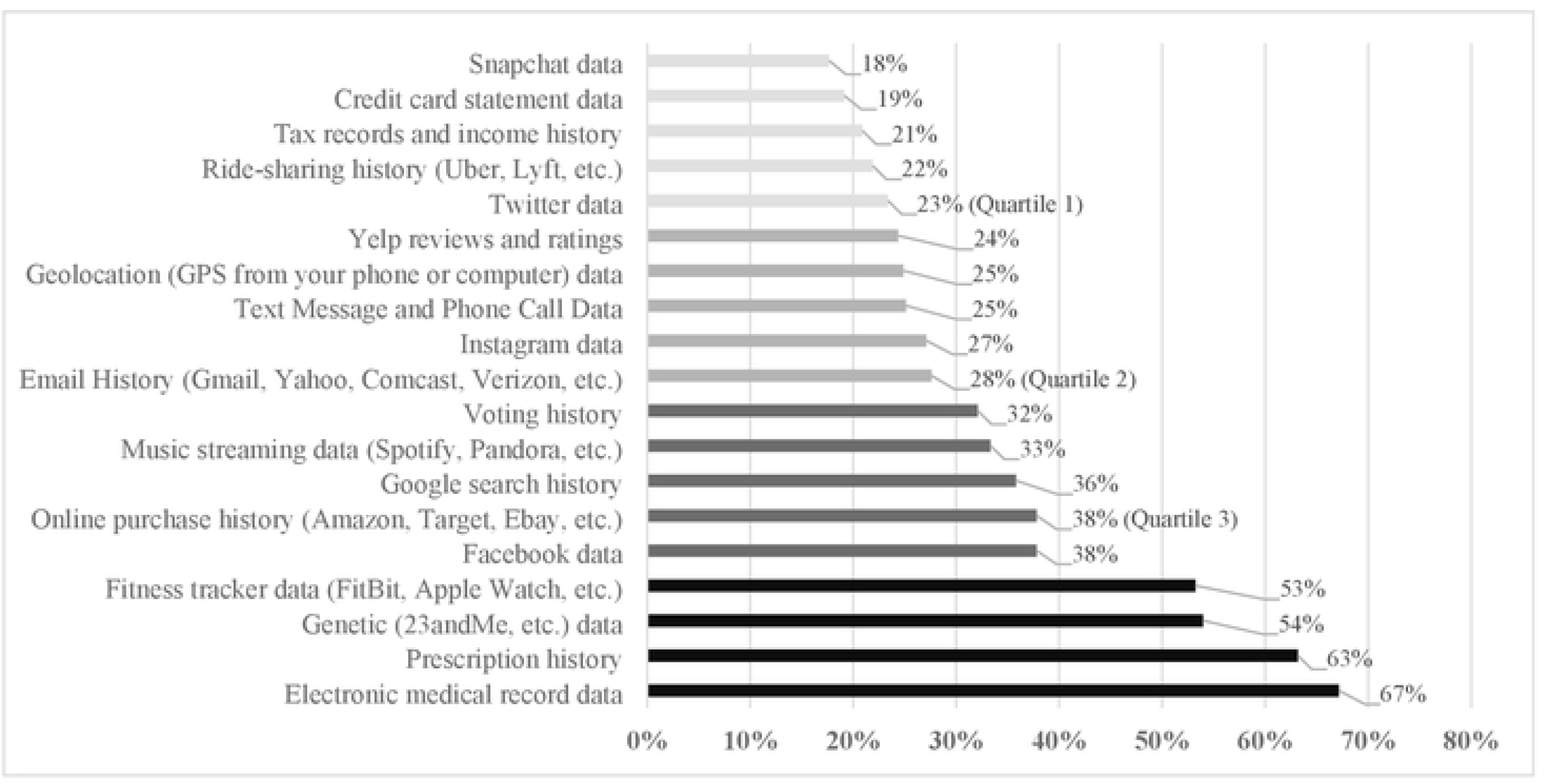
Participant willingness to donate data social media, health, direct communication, online browsing or streaming, financial, location, and voting history data after death (n= 399)

Of people who responded to the demographic questionnaire (n=397), we report our analysis of the relationship between willingness to donate data after death and demographic variables below. Most participants (> 67%) across all demographic responses were willing to donate some of their data after death. Less than 22% of participants across all demographic categories/subcategories were unwilling to donate any of their data and <11% were willing to donate all of their data after death levels (see Table 2 and corresponding Figure 2). Interestingly, all (100%) of participants with an annual frequency of getting ill of 7 to 10 years were willing to donate some of their data after death, comparing to 80% of participants with an annual frequency of getting ill of 4 to 6 years.

**Table 2.** Table summary of participants’ willingness to donate none, some, or all data after death per reported duration of using online medical websites, age range, annual frequency of getting ill, and education level.

| **Willingness to Donate Data After Death** | **Duration of Using Online Medical Websites (Years), N= 397** |  |  |  |  |
| --- | --- | --- | --- | --- | --- |
|  | *Less Than 1* | *2 to 3* | *4 to 5* | *6 to 7* | *More than 7* |
| *None of my data (n= 29)* | 22% | 8% | 9% | 8% | 5% |
| *Some of my data (n= 342)* | 67% | 83% | 79% | 89% | 89% |
| *All of my data (n= 26)* | 11% | 8% | 9% | 3% | 6% |
|  | **Age Range (Years) N= 397** |  |  |  |  |
|  | *30 and under* | *31 to 40* | *41 to 50* | *51 to 60* | *Over 60* |
| *None of my data (n= 29)* | 8% | 12% | 5% | 7% | 6% |
| *Some of my data (n= 342)* | 88% | 82% | 84% | 90% | 84% |
| *All of my data (n= 26)* | 5% | 5% | 9% | 3% | 10% |
|  | **Annual Frequency of Getting Ill, N= 395** |  |  |  |  |
|  | *Less than 1* | *2 to 3* | *4 to 6* | *7 to 10* | *More than 10* |
| *None of my data (n= 28)* | 8% | 5% | 14% | 0% | 0% |
| *Some of my data (n= 341)* | 83% | 89% | 80% | 100% | 94% |
| *All of my data (n= 26)* | 8% | 5% | 6% | 0% | 6% |
|  | **Education Level N= 397** |  |  |  |  |
|  | *High School* | *Some College/*<br>*Associates/ Trade School* | *Bachelors* | *Masters* | *Doctorate or*<br>*other terminal*<br>*degree* |
| *None of my data (n= 29)* | 8% | 5% | 10% | 6% | 6% |
| *Some of my data (n= 342)* | 83% | 88% | 81% | 87% | 91% |
| *All of my data (n= 26)* | 8% | 6% | 8% | 6% | 3% |

**Table 3.** Results from multinomial logistic regression assessing participants’ willingness to donate data after death.

| **Demographic Category** | **Adjusted RRR** | **95% CI** | **P value** |
| --- | --- | --- | --- |
| **Willingness to Donate <u>Some Data</u> versus No Data** |  |  |  |
| Age | 1.10 | 0.84 to 1.43 | 0.49 |
| Education level | 0.95 | 0.63 to 1.43 | 0.81 |
| Duration of using online medical websites | 1.22* | 1.01 to 1.48* | 0.04* |
| Annual frequency of getting ill | 1.16 | 0.73 to 1.85 | 0.52 |
| **Willingness to Donate <u>Some Data</u> versus All Data** |  |  |  |
| Age | 0.84 | 0.63 to 1.11 | 0.22 |
| Education level | 1.14 | 0.76 to 1.72 | 0.53 |
| Duration of using online medical websites | 1.13 | 0.91 to 1.41 | 0.26 |
| Annual frequency of getting ill | 1.18 | 0.74 to 1.90 | 0.50 |
| **Willingness to Donate <u>All Data</u> versus No Data** |  |  |  |
| Age | 1.31 | 0.90 to 1.92 | 0.16 |
| Education level | 0.83 | 0.48 to 1.45 | 0.52 |
| Duration of using online medical websites | 1.08 | 0.82 to 1.42 | 0.59 |
| Annual frequency of getting ill | 0.99 | 0.52 to 1.87 | 0.96 |

\*significant ( $p \leq 0.05$ )

**Figure 3.**
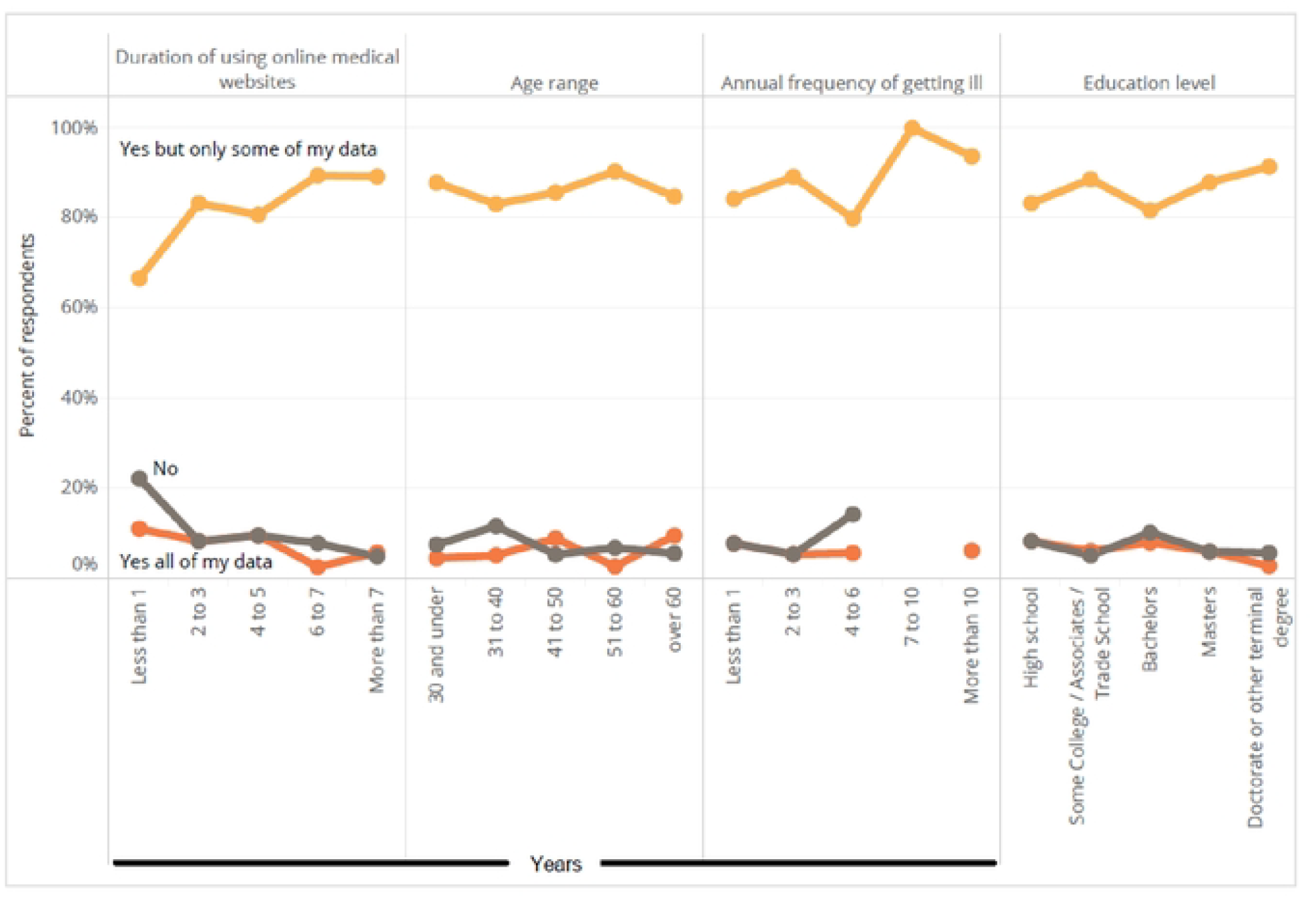
Graphical illustration of participants’ willingness to donate none, some, or all data after death based on duration of using online medical websites (n= 397), age range (n= 397), annual frequency of getting ill (n= 395), and education level (n= 397)

We identified that individuals were more likely to donate some data after death (versus no data) if they had longer duration of using online medical websites (adjusted RRR = 1.22, p<0.05, 95% CI: 1.01 to 1.48), adjusting for age, education, annual frequency of getting ill (see Table 1).

Among individuals willing to donate some data after death and had more than 2 years of using online medical websites, they were more likely to donate health data (electronic medical record data; prescription history data; genetic data, and fitness tracker data; supplement Table 1), similar to the overall participant population (Figure 1). No additional significant findings were observed between willingness to donate all, some, or no data and other demographic variables.

## Discussion

This study is the first to explore a national sample of ResearchMatch participants’ willingness to donate data after death with researchers. This is a timely exploration of adults’ preferences to share data after death with health researchers who are working towards improvement in the standard of care, developing new or optimize current treatments with precision medicine applications, and/or building our generalizable knowledge about health and disease in the real world. Given our finding that individuals who participate in online and/or digital economies are willing to donate some data, and perhaps more so as their duration of using online medical websites increases, individuals could be empowered with tools to communicate and (pre)specify which data they wish to donate and with whom after death. This is especially true and important to help navigate individuals’ personal preferences to share data (or not share data/be forgotten without digital resurrection) after death amid complex privacy and tort laws, research enterprise regulations, and potential family and other surviving caretaker concerns and needs.^19^ Further work might explore whether individuals within online communities might be more willing to donate data to health researchers once the individuals are deceased. Such studies might be useful for researchers who use online mechanisms to engage and educate individuals, prior to and during their engagement in health research, about how their data can be safely processed and/or used upon death during a study.

Although little to no empirical studies have been published on the topic of posthumous data donation, our findings can be compared to a few. For instance, one recent patient survey study, conducted in an emergency room setting, found that most participants (65%; n= 160) expressed willingness to share data from at least one digital data type listed in their survey after death.^20^ This finding is consistent with our finding that most participants (>67%) across all demographic categories were willing to donate some of their data with researchers after death. This same study also found that over 70% of its participants were willing to donate prescription history, electronic medical record, wearables, genetic, and Facebook data.^20^ Their findings largely concur with our present study findings, except we found that less than 50% of participants indicated willingness to donate Facebook data after death (see Figure 1). Therefore, future work might explore the potential impact of surveys conducted among patients or participants within emergency room settings versus online settings to evaluate willingness to donate data after death.

Research and commentary are emerging with discussions on posthumous data donation for secondary research purposes and data use consent strategies. This includes but is not limited to work describing meta-consent versus potentially controversial uses of broad consent, use of data without consent when considered ethically or morally permissible, and ‘contextual exceptionalism’ that would require evaluating uses of data from deceased individuals on a case-by-case basis by research ethics committees.^21–23^ Such work can be useful for practicing researchers and research oversight boards to identify or develop best practices and consent tools, especially in cases where post-mortem data privacy laws might be either nonexistent or vague in covering a wide range of data sources used for research purposes.

For example, commentary highlighting consent models that protect the interests of data subjects, giving them opportunities to expressly opt-out of or opt-in to research using their data upon their death, are useful to develop checklists, forms, and other working documents to support the practice of protecting the interests and autonomy of data subjects. Meta-consent is one such consent model highlighted as a potentially useful alternative in practice to broad consent, a more controversial alternative, for data use after death.^21^ That is, the practice of meta-consent provides data subjects with opportunities to indicate or design how, when, and whether they would like to be approached for data use consent. Under the meta-consent model, individuals could specify research terms, conditions, and contexts (i.e., donation of some, all or no data from one or more data sources to evaluate specific research questions) in which they would consent to the posthumous use, nonuse, or deletion of their personal data.

One recent study that surveyed 100 nonfederal acute care hospital websites to explore whether their privacy policies accurately disclose their use of third-party tracking technologies.^24^ The study found that 96.0% of hospital websites had at least one third-party data request and 86.0% had at least one third-party cookie (n= 100); although, among the 71 accessible privacy policies found only 40 (56.3%) specifically named third-party companies receiving user data.^24^ Therefore, our study, among others, indicates that there are opportunities for health systems, medical journal publishers, and others to determine best practices to safely, creatively, accessibly, and purposefully engage patients in health systems’ online medical websites for educational purposes.^25–28^ Doing so might encourage or inspire individuals to expressly engage in posthumous data donate for health research purposes.

For instance, online medical websites could be developed in ways that might address popular and/or identified reasons for health information seeking behaviors among demographically and geographically diverse populations (i.e., private self-screening, caregiving, learning about clinical trials, creating and/or sharing new information for others, etc.).^28–30^ Owners of websites with moderate to high user activity, or website owners seeking to build towards moderate to high activity, might do so with our present findings in mind. That is, to explore opportunities to develop or embed tools that help prospective study participants connect with researchers and/or opt-in to receiving notifications about opportunities to participate in data-driven research.

Our study is accompanied by general limitations, some of which have been mentioned in our prior publications.^17,18^ For instance, our study sample was derived from ResearchMatch, a platform that uses a meta-consent model to directly reach and engage a population that is likely inclined to engage in data sharing for research. Future work should explore the validity of our key findings among other representative samples of the US population. Also, given that none of our survey respondents with an annual frequency of getting ill at 7 to 10 indicated a willingness to share neither all or none of their data after death, further work could explore this particular sub-demographic to either confirm or gather further qualitative context around this finding.

## Conclusion

Navigating the procedurally complex landscape of post-mortem data privacy and within the data-driven health research landscape warrants the need to explore patterns in individuals’ willingness to donate data with health researchers after death and development of practical recommendations. Our study shows that not only are individuals likely willing to donate health data after death; people are more likely to donate some data after death as they have greater years of using online medical websites. Therefore, our study provides useful insights into how a nationally representative sample of ResearchMatch participants are willing to donate data after death with researchers, which is one route to help guide this complex landscape. Likewise, our findings are useful to those exploring opportunities and strategies to safely and meaningfully engage individuals with online health information seeking behaviors in data-driven health research, both actively and posthumously.

## Data Availability

Data may be made available upon request and according to institutional and ethical guidelines and standards and applicable laws.

## Author Contributions

Conceptualization, R.M.H-S., C.Y.L.; methodology, R.M.H-S., C.Y.L.; formal analysis, R.M.H-S., C.Y.L.; investigation, R.M.H-S., C.Y.L.; data curation, R.M.H-S., C.Y.L.; writing—original draft preparation, R.M.H-S.; writing—review and editing, R.M.H-S., C.Y.L.; visualization, R.M.H-S.; supervision, C.Y.L.; project administration, R.M.H-S., C.Y.L. All authors have read and agreed to the published version of the manuscript.

## Funding

No funding to report.

## Conflicts of Interest

No conflicts of interest to report.

## Acknowledgements

We would like to acknowledge Eric Monson, Ph.D., and Ryan Denniston, Ph.D., within the Duke University Libraries’ Center for Data and Visualization Sciences for advising our project team in our statistical analyses and data visualization processes. We would also like to acknowledge Fang Zhang, Ph.D., at Harvard Pilgrim Health Care Institute for advising our project team in our statistical analyses.

## Notes

### Competing Interest Statement

The authors have declared no competing interest.

### Funding Statement

The author(s) received no specific funding for this work.

